# Tripelennamine 1% topical cream versus diphenhydramine 1% topical cream for the relief of histamine-induced itching: A proof-of-concept study in healthy adults

**DOI:** 10.64898/2026.09.01.26361939

**Authors:** Adwoa O. Nornoo, Harm Maarsingh

## Abstract

**Introduction:** There is an unmet need for effective topical anti-pruritic medications for acute itch, as there are only a few over-the-counter products that have a direct effect on itch. Tripelennamine is a first-generation antihistamine that would be useful in treating histamine-induced pruritus, however, supportive robust clinical data is lacking.

**Objectives:** The efficacy of tripelennamine (TPA) compared to diphenhydramine (DPH) and a vehicle control cream base on histamine-induced pruritus was evaluated as the primary endpoint. Histamine-induced urticaria served as the secondary endpoint.

**Methods:** Thirty-six healthy participants completed this single-center, double-blinded, placebo-controlled crossover clinical study. Following pretreatment with TPA1%, DPH 1% or vehicle control creams, histamine challenge occurred via iontophoresis and a visual analog scale (VAS) for pruritus was used to determine extent of itch (AUC-VAS), peak itch, and duration of itch.

**Results:** Compared to the vehicle control, TPA reduced histamine-induced extent of itch (AUC-VAS), peak itch, and itch duration by 59%, 38% and 43%, respectively (p<0.01 all). DPH did not significantly affect these responses and TPA was superior in reducing extent of itch (48% reduction, p<0.05) and duration (38% shorter, p<0.05). TPA, but not DPH, also reduced histamine induced flare and wheal responses (secondary endpoints) by 53% and 27%, respectively. The reduction in flare responses by TPA was superior to that of DPH (45% reduction, p<0.05).

**Conclusion:** TPA significantly attenuated histamine-induced pruritus and urticaria in a human histamine-challenge model and demonstrated greater efficacy than DPH. These findings provide strong evidence of the antipruritic activity of topical TPA and support further clinical investigation of TPA as a treatment for histaminergic itch and related dermatologic conditions.

## Introduction

Histamine, the main mast cell mediator released during allergic reactions, is responsible for allergy symptoms such as itching, sneezing, runny nose and watery eyes. If histamine is administered intradermally, an inflamed red spot appears within 15 seconds, followed by the appearance of an erythematous flare around the red spot within 1 minute and a light wheal with palpable edema within a few minutes, this is known as the ‘triple response of Lewis’. These responses, including pruritus, can also be induced in non-atopic subjects, making intradermal histamine administration a useful model to explore therapeutic actions of drugs affecting histamine.[1] In the skin, histamine released by skin mast cells and basophils, induces itching, increases skin blood perfusion, and augments microcapillary permeability.[2] Itch results from histaminergic itch transmitted by nociceptive C-fibers, and non-histaminergic itch transmitted by polymodal nociceptive mechano-sensitive C-fibers.[3] Histamine excites fibers expressing H1 and H4 receptors that terminate within and below the stratum granulosum of the skin’s epidermis.

Tripelennamine hydrochloride (TPA), also known as pyribenzamine (PBZ), and diphenhydramine hydrochloride (DPH) are first-generation antihistamines that competitively block peripheral H1 receptors. Both TPA and DPH are lipophilic, aromatic amines, that cross the blood-brain barrier. TPA, like other H1-antagonists, is highly selective for H1 receptors compared with H2, H3, and H4 receptors.[4] In addition, TPA binds and inhibits several other targets, including muscarinic cholinergic receptors, albeit to a lesser extent. The anticholinergic action of DPH is more pronounced, due to a weaker selectivity for H1 receptors over muscarinic receptors.[5] Additionally, both TPA and DPH exert local anesthetic effects[6] through sodium channel blockade.[7]

Limited clinical trials have tested the effectiveness of oral and topical TPA to relieve itching and pain. In a clinical trial without a control group, treatment with TPA 2% cream alleviated pruritus in 80% of subjects with reaction to mosquito bites.[8] In another study, intravenously, orally and topically administered TPA to healthy subjects increased the histamine concentration needed to produce a wheal.[9] TPA ointment was applied on the leg for a systemic effect, whereas the histamine challenge was located on the forearm. In a head-to-head study with oral Benadryl® (DPH) to patients with various skin diseases, TPA had a superior antipruritic effect compared to Benadryl® with fewer side effects.[10] The study was limited by the lack of a vehicle control group, unclear reporting of blinding procedures, and inadequately defined outcome measures, which may affect the interpretability of the findings. TPA and DPH were amongst 5 antihistamines tested for pollen/mold-induced responses, with TPA having more favorable outcomes than DPH. Regarding wheal suppression, TPA ranked 3 and DPH 5 ; for duration of suppression, TPA ranked 2 and DPH 5 ; TPA ranked 2 and DPH 4 regarding the least amount of side effects.[11]

From the forgoing discussion, there is still an unmet need for effective topical medications for acute itch. Our study determined the effectiveness of tripelennamine 1% topical cream compared to diphenhydramine 1% topical cream to relieve histamine-induced itching using a visual analogue itch score (VAS) for pruritus. The antihistamines and the vehicle control cream base were applied to the skin prior to a challenge with histamine delivered via iontophoresis. TPA was hypothesized to have a significantly lower sensation of itching induced by histamine when compared to diphenhydramine or vehicle control cream base as measured by VAS.

## Methods

### Study Formulations

Histamine 1% topical gel was formulated by the principal investigator. A ‘Master Compounding Formulation Record’ was completed each time the gel was made. Histamine (1% w/w) was prepared by combining 0.5g of histamine dihydrochloride 99% (Acros Organics), 1.25g of Methocel® K100M (PCCA), approximately 48mL of 0.9% Sodium Chloride Injection, USP (B Braun) and mixing in a glass beaker till the gel was formed. Tripelennamine 1% topical cream was supplied by Kingsway Pharmaceuticals. Diphenhydramine 1% topical cream and the vehicle control cream base were purchased from local pharmacies.

### Study Design

The primary endpoint of this single center, double-blinded, vehicle-controlled randomized crossover study in healthy subjects was to determine the effectiveness of TPA compared to DPH in relieving histamine-induced pruritus. All research involving human subjects was conducted under the auspices of the Palm Beach Atlantic University, West Palm Beach, FL and received prior approval from its’ Institutional Review Board (IRB).

The study enrolled healthy adults aged 18–50 years from Palm Beach Atlantic University, West Plam Beach, FL, USA, with no history of chronic itch or pain, and avoided products that could affect itch or pain perception for one week. Participants were excluded if they had skin conditions or diseases affecting itch or pain perception, used interfering emollients, were enrolled in another investigational study, had allergies to the product excipients, or were pregnant. Subjects were enrolled in the study after completing and signing an informed consent form.

The VAS served as the primary endpoint to assess itch induced by the iontophoretically delivered histamine. It is a validated test for the assessment of pruritus and the potential effectiveness of treatments, such as antihistamines.[3, 12] The following parameters were determined from the mean VAS vs time curve: area-under-the-curve (signifying extent of itch experienced), duration of itch, and peak itch (highest VAS recorded). Subjects who satisfied the inclusion and exclusion criteria were assigned a unique randomized Subject ID number upon enrollment (Qualtrics, Provo, UT. Four 4x4 cm test areas were demarcated on the volar forearms, 2 on each arm, each separated by 4 cm. The creams (0.2g) were applied in a double-blinded manner to the demarcated skin areas using a spatula. The vehicle control cream base (CB) served as the control for each of the antihistamine creams, hence were placed on both arms. Two blinded sets (TPA and CB, DPH and CB) were randomly assigned by a second investigator, and the blinded treatment codes were recorded with the Subject ID number.

Fifteen minutes after application, any remaining cream was gently wiped off, and histamine was applied iontophoretically to the site, in line with previous studies.[13] Acute histaminergic itch was induced by placing an anode delivery electrode (Trivarion®, North Coast Medical, Inc; pre-soaked with 0.2 mL normal saline) containing 0.2g of histamine 1% topical gel in the center of the test area, while a cathode grounding electrode was placed on the subject’s arm 4 cm outside the test area. The iontophoresis device (ActivaDose II Device, Activa Tek, Taiwan) generated a current of no more than 0.5 mA at a dose of 1.0 mA x min. One area was tested at a time to allow for adequate VAS evaluation. Treatments to the next area were not applied until the VAS score returned to zero on the test area.

Subjects rated their perception of itch every 60 seconds by pointing to a horizontal VAS scale (Figure 1) until the itch sensation had completely subsided. Scores were recorded by a research assistant supervised by the blinded investigator. VAS values were rated as follows: 0 – no pruritus, 1-3 - mild pruritus, 4-6 - moderate pruritus, 7-8 - severe pruritus and 9 - very severe pruritus. Secondary endpoints included the development of urticaria: wheal and flare magnitude, which were evaluated and scored every 5 minutes for up to 20 minutes: 1- no/mild, 2- moderate and 3- severe. A medical director was present onsite, and participants were instructed to report any adverse events up to one week after study completion.

**Fig. 1.**
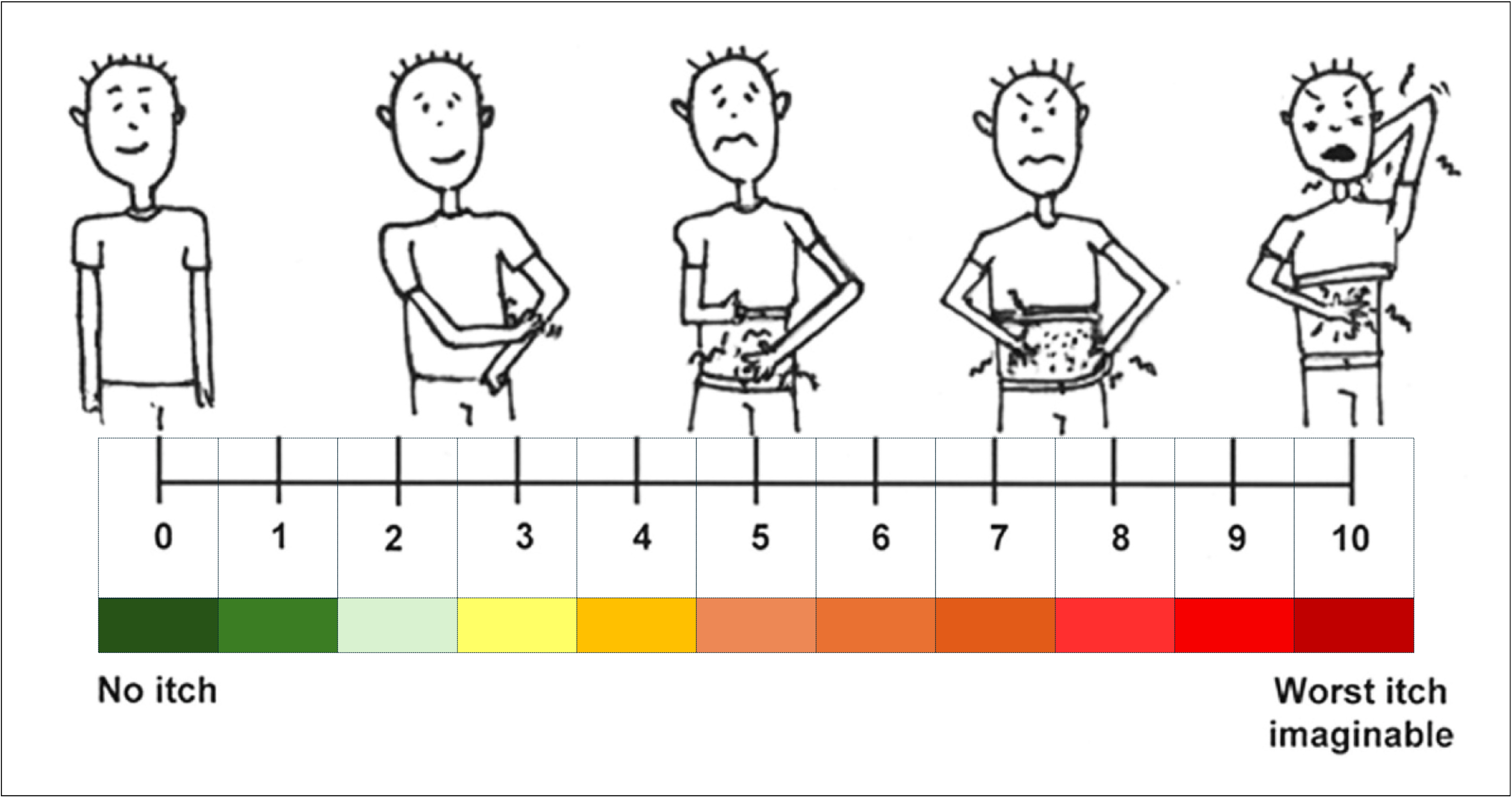
Visual Analog Itch Score (VAS) scale. Adapted from Haydek, et al.[12]

### Statistical analysis

The following parameters were evaluated: area-under the-curve (AUC-VAS), peak VAS, duration of itch, flare magnitude and wheal magnitude. Power analysis revealed that at a probability of p<0.05 and a power of 0.95, 36 participants were needed for statistical inference. Normality assessment was tested by Sharpiro-Wilks tests. Endpoints were compared statistically amongst the three treatments using a repeated measures Friedman’s ANOVA, Kruskall-Wallis ANOVA, and Fisher Exact tests, as appropriate, with Dunn’s post-hoc analysis for individual differences at p<0.05 (OriginPro, Version 2026. OriginLab Corporation, Northampton, MA, USA).

## Results

Participants were recruited through IRB-approved flyers over a 6-month period. Thirty-seven participants were recruited (Table 1). Of the 36 participants who completed the study, 73% were females, White participants comprising most of the cohort. Most participants were young adults between 19 and 30 years of age reflecting the pool from which the participants were drawn. One participant experienced a mild local skin reaction attributed to an excipient in the TPA cream and withdrew from the study before completion. The event resolved without intervention and without sequelae. No other treatment-related adverse events or clinically significant local or systemic adverse effects were reported.

**Table 1.** Study Participant Demographics.

| **Table 1. Study Participant Demographics** |  |  |
| --- | --- | --- |
|  | **Female** | **Male** |
| Number of Participants | 26 | 10 |
| **RACE** |  |  |
| Asian | 0 | 1 |
| Black/African American | 6 | 2 |
| Other | 1 | 1 |
| White | 20 | 5 |
| **AGE (Years)** |  |  |
| 19-30 | 24 | 6 |
| 31-50 | 4 | 2 |
| Mean (SEM) | 25 (1) | 35.7 (1.9) |
| Minimum | 19 | 22 |
| Maximum | 48 | 50 |
| **BMI (kg/m<sup>2</sup>)** |  |  |
| Mean (SEM) | 25.2 (1.1) | 25.01 (0.5) |
| Minimum | 14.8 | 20.3 |
| Maximum | 48.9 | 29.7 |

### Primary Endpoints

The effectiveness of TPA compared to DPH in relieving histamine-induced itching using a VAS assessment was determined. For CB treated areas, the extent of histamine-induced itch increased with time reaching a distinct peak and declining thereafter (Figure 2). This was also observed in the DPH-treated group, but not the TPA-treated group where the decline occurred immediately. AUC-VAS, a measure of the extent of itch experienced, was calculated from the VAS versus time plot for each treatment (Figure 3). Mean AUC-VAS values were 32.4±4.7, 25.7±4.0, and 13.3±2.8 for CB, DPH and TPA, respectively. AUC-VAS for TPA was 59% (p<0.01) and 48% (p<0.05) lower compared to CB and DPH, respectively, a ∼2.5-fold and 2-fold lower extent of itch experienced. This indicates the ability of TPA to prevent the effects of histamine. In contrast, DPH did not significantly reduce the AUC-VAS.

**Fig. 2.**
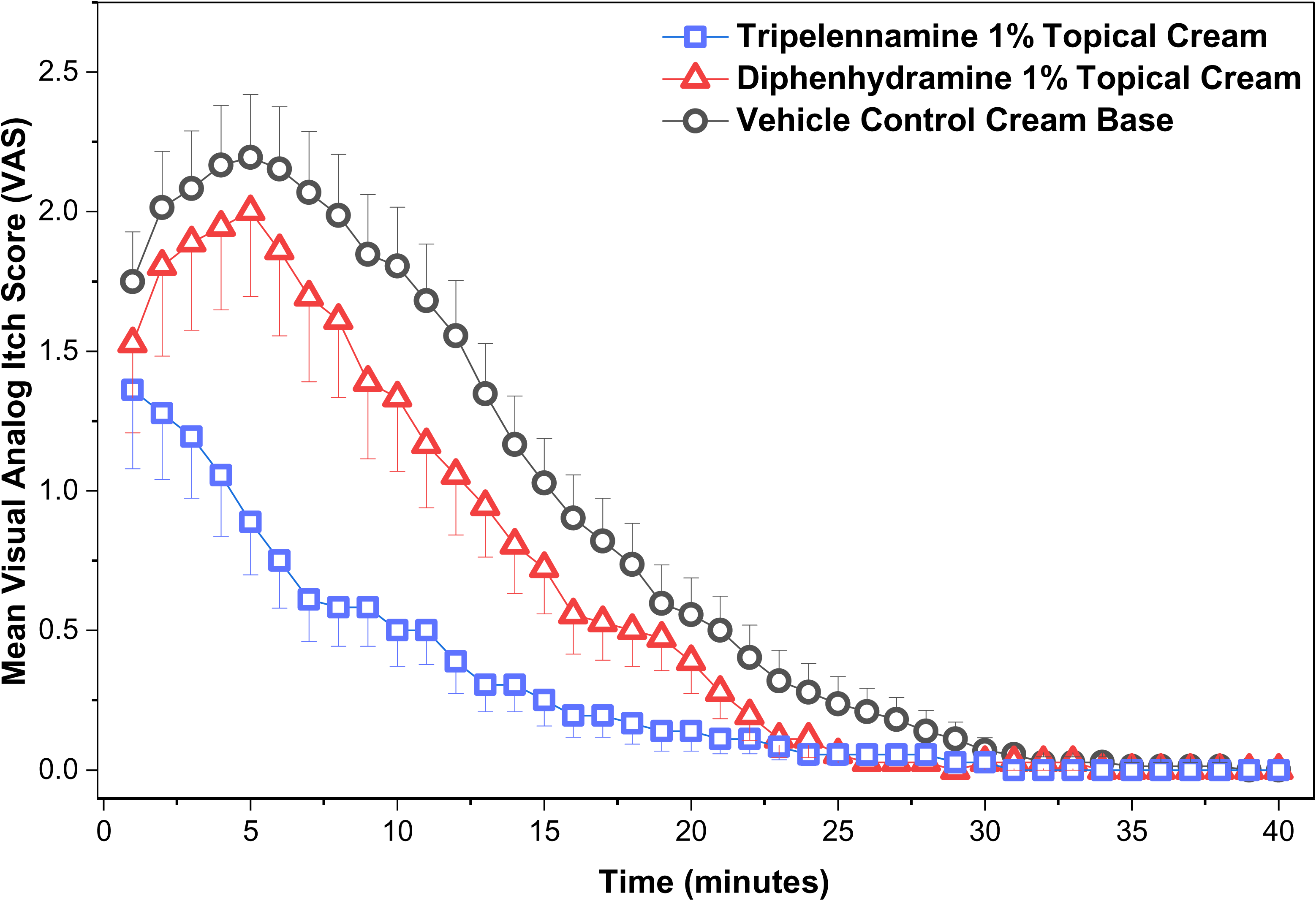
Mean Visual Analog Itch Score (VAS) versus time for CB, TPA and DPH. Mean±SEM of 36 participants.

**Fig. 3.**
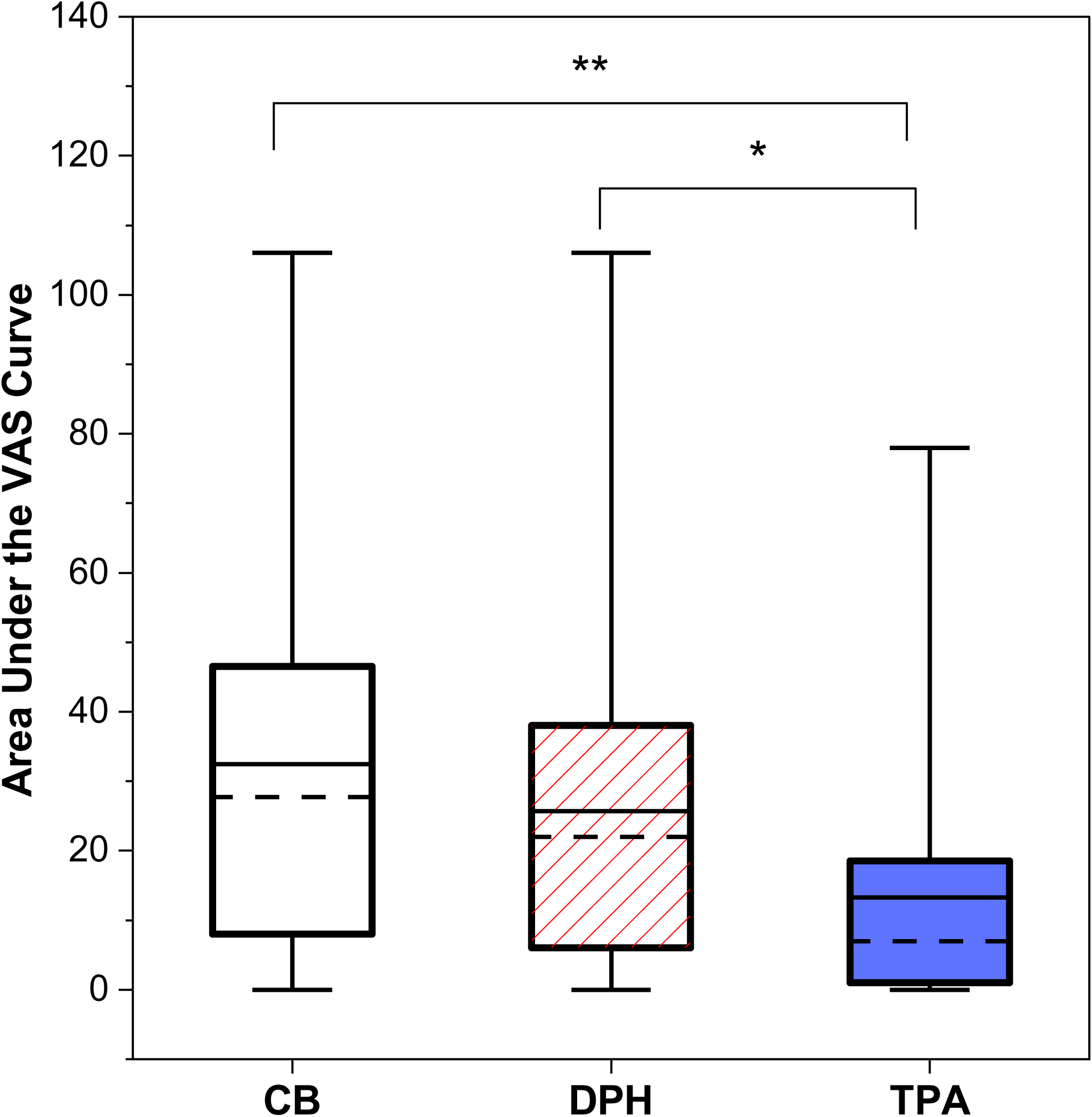
Area under the VAS curve for CB, TPA and DPH. Box plots illustrate median (dashed line) and mean (solid line); interquartile range (IQR; 25th–75th percentiles), and minimum and maximum values. **p<0.01, *p<0.05 compared to TPA.

The peak itch was recorded as the highest VAS following the histamine challenge. The lower the peak VAS the greater the attenuation of the effects of histamine. Peak VAS was 2.8±0.3, 2.4±0.3, and 1.7±0.3 for CB, DPH and TPA, respectively (Figure 4, Table 2). The reduction in peak itch by TPA was significant compared to CB (38% reduction, p<0.01) but not compared to DPH. The mean peak for CB indicates an overall mild itch sensation following the histamine challenge. Treatment with TPA resulted in a lower number of participants experiencing moderate-to-severe itch (p<0.05) and more experiencing no itch (p<0.05) compared to CB (Table 2). DPH did not affect mean peak VAS.

**Fig. 4.**
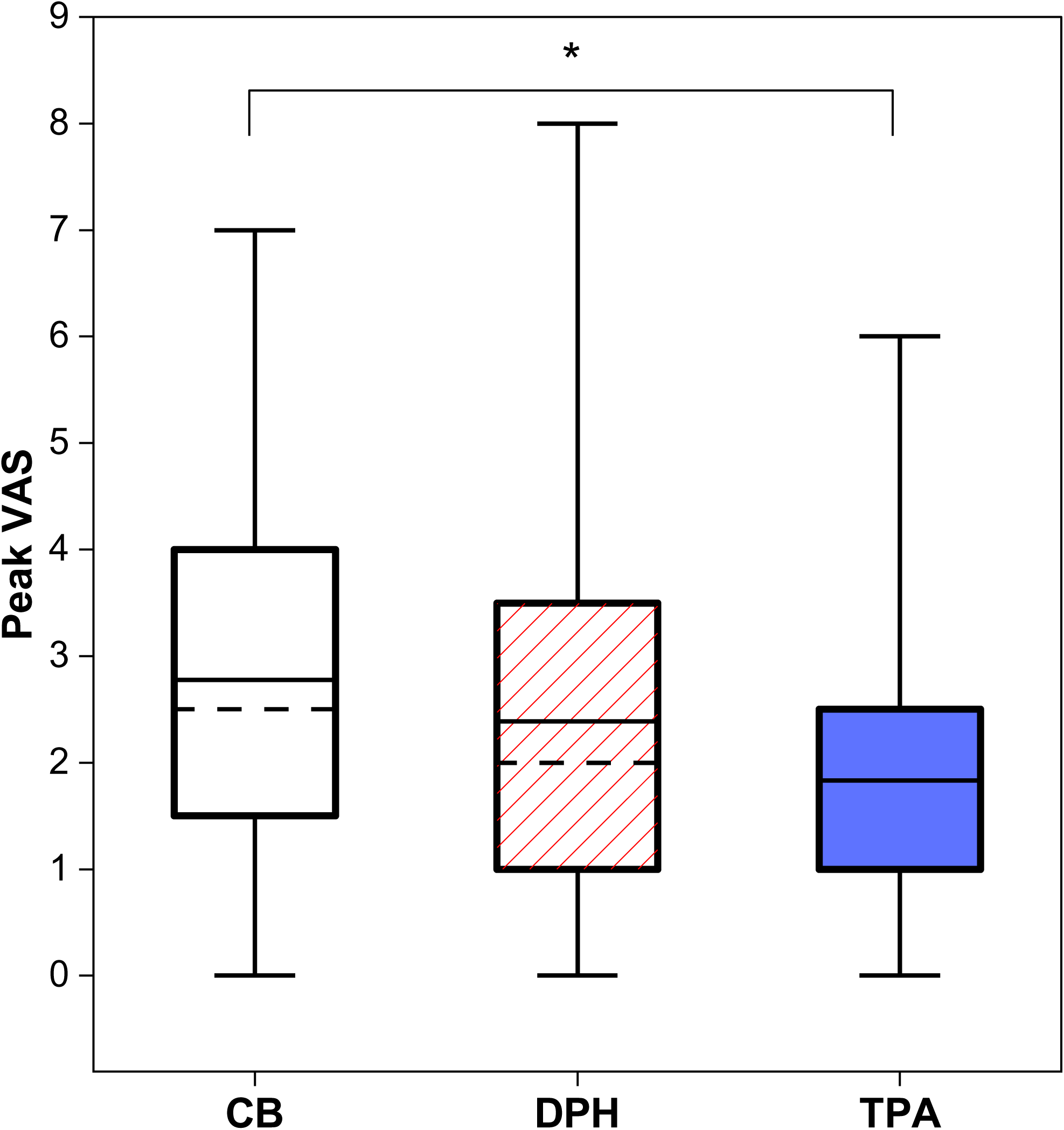
Peak VAS for CB, TPA and DPH. Box plots illustrate median (dashed line) and mean (solid line); interquartile range (IQR; 25th–75th percentiles), and minimum and maximum values. Median dashed line overlaps the 25th percentile for TPA. *p<0.01 compared to TPA.

**Table 2.** Classification of Study Participant VAS.

| Table 2. Classification of Study Participant VAS |  |  |  |  |  |
| --- | --- | --- | --- | --- | --- |
|  | Mean Peak VAS | No itch<br>VAS 0 | Mild itch<br>VAS 1-3 | Moderate itch<br>VAS 4-6 | Severe itch<br>VAS 7-9 |
| **CB** | 2.8±0.3 | 2 | 20 | 13 | 1 |
| **DPH** | 2.4±0.3 | 7 | 20 | 0 | 2 |
| **TPA** | 1.7±0.3** | 9* | 23 | 4* | 0 |
| *p<0.05, **p<0.01 compared to CB. |  |  |  |  |  |

Duration of itch was determined by recording the last time point a participant reported an itch sensation. Mean itch duration after histamine application was 13.5±1.4 minutes, 12.5±1.4 minutes, and 7.8±1.4 minutes for CB, DPH and TPA, respectively (Figure 5). Itch duration was significantly shorter with TPA compared to either CB (44% shorter, p<0.01) or DPH (38% shorter, p<0.05). In absolute terms, itch duration was reduced by ∼6 and 5 minutes compared with CB and DPH, respectively. DPH did not affect itch duration.

**Fig. 5.**
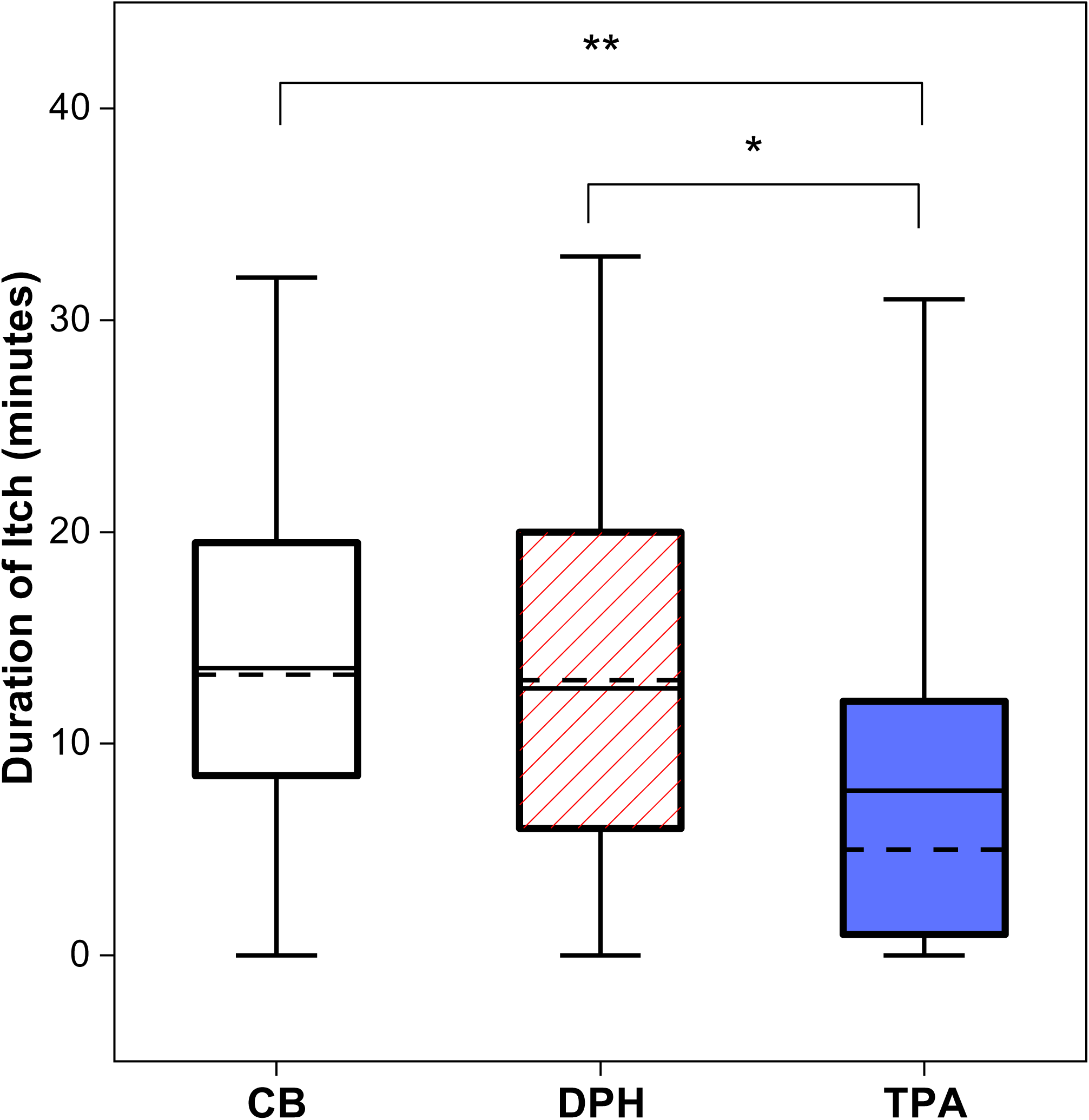
Duration of itch for CB, TPA and DPH. Box plots illustrate median (dashed line) and mean (solid line); interquartile range (IQR; 25th–75th percentiles), and minimum and maximum values. **p<0.01, *p<0.05 compared to TPA.

### Secondary Endpoints

The magnitude of a wheal and flare after a histamine challenge, indicative of urticaria, served as secondary endpoints. Histamine induced flare and wheal reactions on the skin after pre-treatment with CB, DPH and TPA to varying degrees (Figure 6). Flare scores were lower with TPA at all time points compared to CB and up to 10 minutes compared to DPH. Mean flare scores were 2.18±0.12, 2.01±0.13, and 1.56±0.11, for CB, DPH and TPA, respectively (Figure 7). Mean wheal scores were 2.54±0.09, 2.41±0.12, and 2.23±0.14 for CB, DPH and TPA, respectively. Lower flare (50% reduction; p<0.01) and wheal (27% reduction, p<0.05) responses with TPA relative to CB reflects the positive effects of TPA in preventing urticaria. Pretreatment with DPH on the other hand did not significantly reduce wheal or flare responses. TPA was significantly more effective than DPH in suppressing histamine-induced flare (45% reduction, p<0.05) responses.

**Fig. 6.**
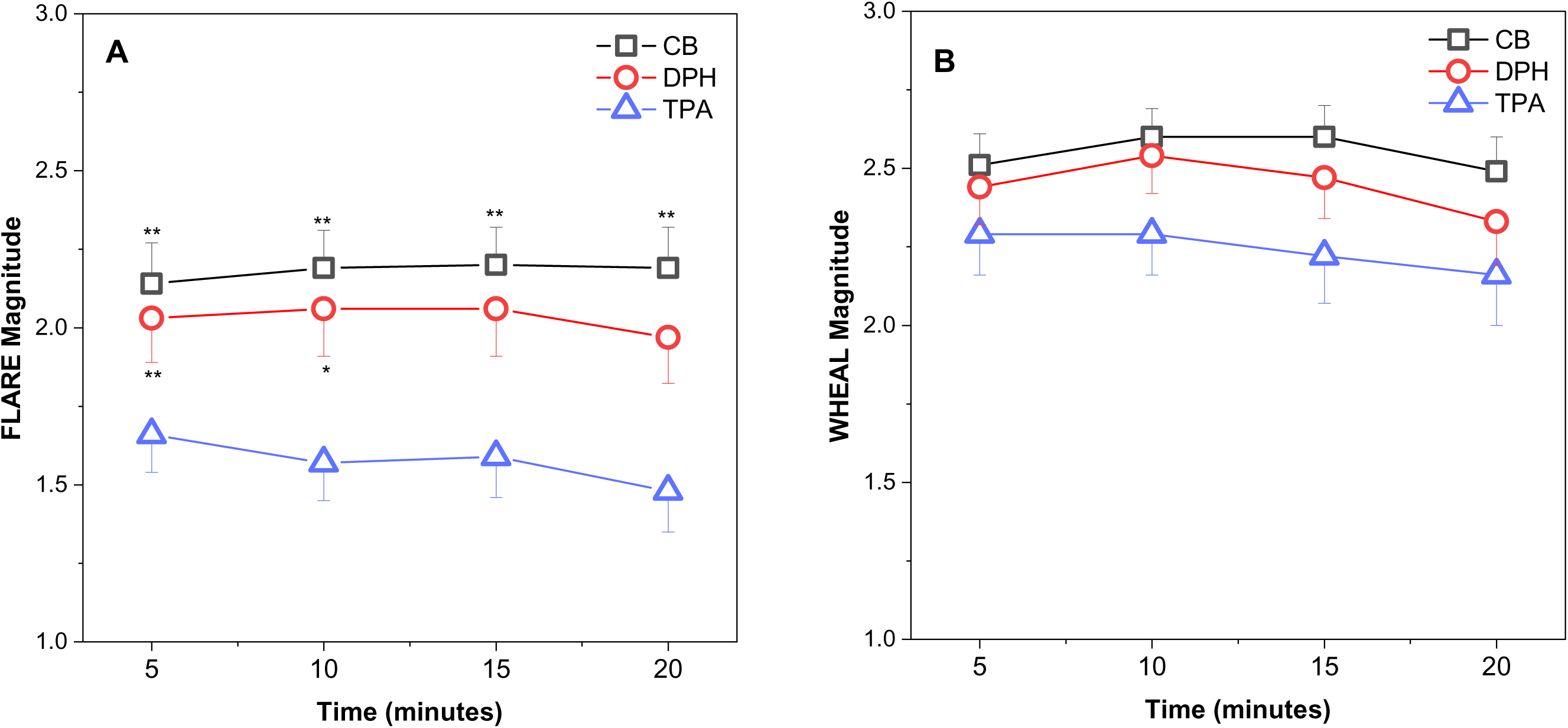
Flare (A) and wheal (B) magnitude over time to histamine challenge for CB, TPA and DPH. Mean±SEM of 36 participants. **p<0.01, *p<0.05 compared to TPA.

**Fig. 7.**
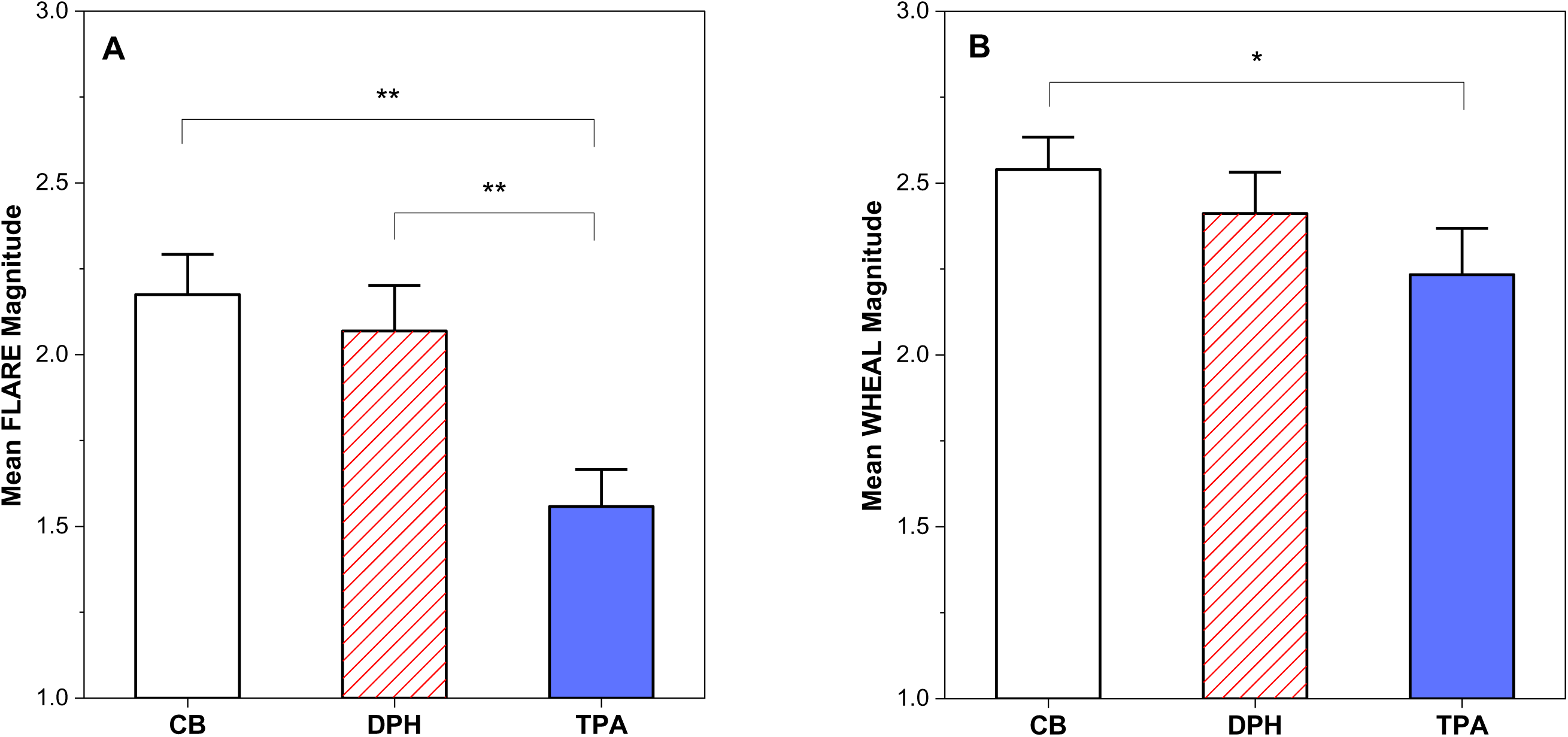
Mean flare (A) and wheal (B) responses to histamine challenge for CB, TPA and DPH. Mean±SEM of 36 participants. **p<0.01, *p<0.05 compared to TPA.

## Discussion

The primary endpoint of this randomized, double-blind, vehicle-controlled crossover human study was to determine the effectiveness of tripelennamine 1% topical cream (TPA) compared to diphenhydramine 1% topical cream (DPH) in relieving histamine-induced pruritus. Compared with the vehicle control (CB), TPA significantly attenuated histamine-induced pruritus, decreasing overall itch sensation, itch duration, and peak VAS scores by 59%, 43%, and 38%, respectively. The secondary endpoint in this study determined the effectiveness of TPA compared to DPH in relieving histamine-induced urticaria. TPA reduced flare and to a lesser extent wheal development. In contrast to TPA, DPH did not significantly reduce pruritic symptoms or urticarial responses. Taken together the results indicate antipruritic and anti-urticarial activity of TPA in a validated human histamine-challenge model.

We are unaware of any previous studies demonstrating the antipruritic and anti-urticaria efficacy of TPA against histamine-induced itch. An early study showed that 72% of patients experiencing pruritus reported “consistent relief” after topical application of TPA 2%.[14] In another study, topical TPA 2% was effective in treating pruritus with 82% of participants reporting pruritus as absent or relieved to skin reactions induced by mosquito bites.[8] The antipruritic effect of TPA was very similar to that of the local anesthetic dibucaine 1%, therefore, the authors concluded that at least part of the antipruritic effect of TPA could be attributed to its’ local anesthetic effect,[8] a known feature of antihistamines.[6, 7] In a third study, TPA administered intravenously, orally and transdermally to healthy subjects increased the concentration of histamine needed to produce a wheal response.[9] All three studies were limited by the absence of placebo controls, lack of blinding, poorly defined outcomes measures or administration routes that did not directly assess local skin effects.

The observed effects of TPA are consistent with inhibition of histamine-mediated signaling in the skin. Histamine activates peripheral sensory nerve endings through H1 receptors, resulting in itch perception, while also inducing vasodilation and increased vascular permeability that contribute to flare and wheal formation.[1, 2] The reduction in both itch and urticarial responses observed in this study suggests effective suppression of histamine-driven neurovascular signaling. In addition to H1 receptor antagonism, first-generation antihistamines possess local anesthetic properties through sodium channel blockade.[6, 7] Both H1-receptor blocking properties and local anesthetic actions could explain both the antipruritic effects and the reduction in flare observed in our study.[15] These actions may further contribute to attenuation of sensory nerve activation and itch perception. The relative contributions of H1 receptor antagonism and local anesthetic effects were not assessed in the present study and warrant further investigation.

In contrast to TPA, topical DPH treatment did not significantly affect histamine-induced pruritus or urticaria in our study. An early study [16] demonstrated that topical DPH 2% or vehicle control did not affect histamine-induced wheal or flare responses. Moreover, the antipruritic effects of topical DPH in patients with various forms of dermatitis were comparable to those of the vehicle control, with no statistically significant differences observed. The author postulated that the absence of a clinical effect was due to low DPH absorption, even though this was not assessed.[17] In another study, topical application of a higher dose of DPH (5%) increased the histamine concentration needed to induce pruritus by 2.3-fold.[17] In a third study, Philip[18] found that 73% of participants with various forms of itch dermatitis had full or partial relief after long-term treatment with DPH 2%. In more recent studies, topical DPH 2% did not significantly affect peak VAS scores, itch duration, or itch extent in cowhage-induced pruritus, although in a study lacking a vehicle control, subjects with epidermolysis bullosa reported that topical DPH “relieved itch a little.” [19–21] Although topical DPH remains widely available as an over-the-counter antipruritic product, robust evidence supporting its effectiveness is limited.[22] The present study further suggests that TPA may provide greater suppression of histamine-mediated skin responses than DPH when administered topically.

Acute histaminergic itch contributes to symptoms associated with insect bites, allergic skin reactions, urticaria, and minor skin irritation. The magnitude of itch suppression observed with TPA in the present study suggests that topical TPA may represent a useful non-steroidal treatment option for such conditions. The reduction in both sensory (itch) and vascular (flare and wheal) responses further supports its potential therapeutic utility. However, clinical studies in patients with pruritic dermatologic disorders are needed to determine whether the benefits observed in this experimental model translate to clinical practice.

The strength of this study is that it is a double-blinded, placebo-controlled, crossover clinical study that was sufficiently powered. The application of the creams 15 minutes prior to a histamine challenge[2] reduced confounding factors, such as a potential sense of relief by the application process. Application prior to the histamine challenge would make assessing the onset of action difficult. Since the sensation of itch is subjective, proper instruction on obtaining a VAS was provided. Moreover, the crossover design of the study reduced subjectivity between treatment groups.

Several limitations, however, should be acknowledged. First, the study was conducted in healthy subjects instead of patients with dermatologic disorders. Second, histamine-induced itch is an experimental model of acute histaminergic pruritus and may not fully represent chronic itch conditions in which non-histaminergic mechanisms contribute substantially to symptom generation. Third, the study evaluated short-term responses following a single application and therefore does not address long-term effectiveness, safety, or tolerability. Future studies should evaluate TPA in patients with clinically relevant pruritic disorders and determine its effectiveness under real-world treatment conditions.

In conclusion, topical TPA significantly attenuated histamine-induced pruritus and urticaria in a human histamine-challenge model and demonstrated greater efficacy than topical DPH. These findings provide strong evidence of the antipruritic activity of topical TPA and support further clinical investigation of TPA as a treatment for histaminergic itch and related dermatologic conditions.

## Acknowledgement

The authors thank the medical director Dr. Jennifer Fleigelman and the following research assistants for their role in conducting the study: Adrian Almeida, McKenna Brown, Hugh Buera, Madison Dong, Mariah Gonzalez, Dana Klassen, Melanie Lafita, Katherine Lui-Golian and Brianna Velardi.

## Statement of Ethics

### Study approval statement

The study protocol was reviewed and approved by the Institutional Review Board of Palm Beach Atlantic University (Protocol No. IIIc 01.21.2025). The study was conducted in accordance with the ethical principles outlined in the Declaration of Helsinki and applicable institutional guidelines for research involving human subjects. The study was retrospectively registered at ClinicalTrials.gov (Identifier: NCT07778472). At the time the study was conducted, prospective clinical trial registration was not required by the Institutional Review Board for this type of investigator-initiated healthy-volunteer research. To promote transparency and public accessibility of the study information, the protocol was subsequently registered retrospectively.

### Consent to participate statement

Written informed consent was obtained from participants to participate in this study. Approval of the consent form was obtained from the Institutional Review Board of Palm Beach Atlantic University before any participant was enrolled.

## Author Contributions

Adwoa O. Nornoo, PhD, Principal Investigator, was responsible for the design, performance, data analysis, scientific reporting and professional direction of this project.

Harm Maarsingh, PhD, Co-Principal Investigator was responsible for conducting the clinical trial, data analysis and scientific reporting.

## Data Availability Statement

All data generated or analyzed during this study are included in this article. Further enquiries can be directed to the corresponding author.

## References

1. Thurmond RL, Kazerouni K, Chaplan SR, Greenspan AJ. Peripheral Neuronal Mechanism of Itch: Histamine and Itch. In: Carstens E, Akiyama T, editors. Itch: Mechanisms and Treatment. Frontiers in Neuroscience. Boca Raton (FL): CRC Press, Taylor and Francis; 2014.

2. Heinemann C, Elsner P. Efficacy measurement of topical antihistamines: a review. Skin Pharmacol Appl Skin Physiol. 2003;16(1):4–11.

3. Andersen HH, Sorensen AR, Nielsen GA, Molgaard MS, Stilling P, Boudreau SA, et al. A Test-Retest Reliability Study of Human Experimental Models of Histaminergic and Non-histaminergic Itch. Acta Derm Venereol. 2017;97(2):198–207.

4. BindingDB [Internet]. University of California, San Diego. 2024 [cited July 9, 2026]. Available from: https://www.bindingdb.org.

5. Kubo N, Shirakawa O, Kuno T, Tanaka C. Antimuscarinic effects of antihistamines: quantitative evaluation by receptor-binding assay. Jpn J Pharmacol. 1987;43(3):277–82.

6. Rumore MM, Schlichting DA. Analgesic effects of antihistaminics. Life Sci. 1985;36(5):403–16.

7. Kuo CC, Huang RC, Lou BS. Inhibition of Na(+) current by diphenhydramine and other diphenyl compounds: molecular determinants of selective binding to the inactivated channels. Mol Pharmacol. 2000;57(1):135–43.

8. O’Rourke FJ, Murnaghan MF. The cutaneous reaction to the bite of the mosquito Aedes aegypti (L.) and its alleviation by the topical application of an antihistaminic cream (pyribenzamine). J Allergy. 1953;24(2):120–5.

9. LeVan P, Sternberg TH, Perry DJ. Studies of the “antihistaminic” effect of pyribenzamine administered by various routes. Calif Med. 1951;74(4):256–9.

10. Morrow G. Benadryl and Pyribenzamine in the Treatment of Diseases of the Skin. Calif Med. 1948;69(1):22–4.

11. Cook TJ, MacQueen DM, Wittig HJ, Thornby JI, Lantos RL, Virtue CM. Degree and duration of skin test suppression and side effects with antihistamines. A double blind controlled study with five antihistamines. J Allergy Clin Immunol. 1973;51(2):71–7.

12. Haydek CG, Love E, Mollanazar NK, Valdes Rodriguez R, Lee H, Yosipovitch G, et al. Validation and Banding of the ItchyQuant: A Self-Report Itch Severity Scale. J Invest Dermatol. 2017;137(1):57– 61.

13. Balogun JA, Adeniyi EA, Akala EO. Cardiovascular responses during histamine iontophoresis therapy. Aust J Physiother. 1991;37(2):105–10.

14. Feinberg SM, Bernstein TB. Tripelennamine pyribenzamine ointment for the relief of itching. J Am Med Assoc. 1947;134(10):874.

15. Rosa AC, Fantozzi R. The role of histamine in neurogenic inflammation. Br J Pharmacol. 2013;170(1):38–45.

16. Perry DJ. The local use of benadryl ointment. J Invest Dermatol. 1947;9(2):95–7.

17. Bernstein JE, Whitney DH, Soltani K. Inhibition of histamine-induced pruritus by topical tricyclic antidepressants. J Am Acad Dermatol. 1981;5(5):582–5.

18. Philip AJ. Treatment of itching dermatoses with an ointment containing 2% diphenhydramine (benadryl hydrochloride). N Y State J Med. 1949;49(10):1179–81.

19. Papoiu AD, Chaudhry H, Hayes EC, Chan YH, Herbst KD. TriCalm(®) hydrogel is significantly superior to 2% diphenhydramine and 1% hydrocortisone in reducing the peak intensity, duration, and overall magnitude of cowhage-induced itch. Clin Cosmet Investig Dermatol. 2015;8:223–9.

20. Papoiu AD, Valdes-Rodriguez R, Nattkemper LA, Chan YH, Hahn GS, Yosipovitch G. A novel topical formulation containing strontium chloride significantly reduces the intensity and duration of cowhage-induced itch. Acta Derm Venereol. 2013;93(5):520–6.

21. Danial C, Adeduntan R, Gorell ES, Lucky AW, Paller AS, Bruckner AL, et al. Evaluation of Treatments for Pruritus in Epidermolysis Bullosa. Pediatr Dermatol. 2015;32(5):628–34.

22. Eschler DC, Klein PA. An evidence-based review of the efficacy of topical antihistamines in the relief of pruritus. J Drugs Dermatol. 2010;9(8):992–7.

